# The development and usability testing of six arts-based knowledge translation tools for parents about COVID-19

**DOI:** 10.1101/2025.03.31.25324968

**Authors:** Shannon D. Scott, Sarah A. Elliott, Kathy Reid, Lisa Knisley, Lisa Hartling

## Abstract

COVID-19 was declared a global pandemic in March 2020. This novel disease impacted how health information was communicated as information on both the disease itself and on public health guidelines changed rapidly.

The purpose of this research was to create knowledge translation (KT) tools about COVID-19 to increase public confidence in science and to encourage vaccine uptake and maintenance of public health measures. Our goal was to develop, evaluate and disseminate innovative KT tools to increase awareness, knowledge and uptake of evidence about COVID-19 among parents and families. The project had two main sources of data collection: 1) qualitative semi-structured interviews with 27 parents whose children had COVID-19 between May 2020 and April 2022 and 2) focus group discussions with 67 parents between October and December 2021 to understand their experiences and information needs related to COVID-19 public health measures, including vaccination, mask wearing, social distancing, and other public health measures.

Based on the qualitative findings from semi-structured interviews of parents whose children had COVID-19, we developed two KT tool prototypes: a video and an interactive infographic addressing the management of a child with COVID-19. The qualitative findings from focus groups were used to develop 4 KT tool prototypes on 2 topics: COVID-19 vaccines for children and navigating a child’s social world during the COVID-19 pandemic and beyond. Usability testing of all 6 KT tools was completed by parents from our established networks. The tools were revised based on usability results, and final versions of the tools were made publicly available on our website (echokt.ca) in November 2022. We further disseminated the resources through our social media channels and other established stakeholder networks.

## Introduction

COVID-19 emerged as a novel viral disease in late 2019 and by March 2020 was declared a worldwide pandemic (1). Little was known about the virus, including how it spread and who would be most susceptible to severe infection (1). In early phases (or waves) of the disease, children were not becoming as severely ill as adults (2-10). Yet the global response to this pandemic impacted children and families, as schools and activities for children were shut down to all in-person contact. Children were quickly isolated from their peers (11-13).

As the disease continued to evolve, the changing symptoms, diagnostic uncertainty, social isolation and the rollout of new vaccines led to a rapidly changing body of scientific evidence that resulted in uncertainty of how to best manage the impacts on children and families (14-18). There was a critical need to understand information needs and to effectively communicate new and evolving evidence to ensure public safety.

The purpose of this research was to work with parents to develop, refine, evaluate, and disseminate knowledge translation (KT) tools for parents and families to increase their knowledge about the prevention and management of COVID-19, including risk factors for severe outcomes, measures to prevent transmission (vaccination, masks, handwashing), and evidence about safety and efficacy of COVID-19 vaccines.

## Methods

A multi-method study involving patient engagement was used to develop, refine, and evaluate three animation videos and three interactive infographics for parents about managing their children during the COVID-19 pandemic. Parents from the Pediatric Parent Advisory Group (P-PAG) and the Pediatric Parent Consultation Network (P-PCN) were involved throughout the research phases. These two networks are unique to our research program, and facilitate patient engagement throughout the development of KT tools. Members of the P-PAG and P-PCN contribute to research design, tool development, tool evaluation, and dissemination of our research (19, 20). (20) Research ethics approval was obtained from the University of Alberta Health Research Ethics Board (Edmonton, AB) [Pro0062904].

### Compilation of Parents’ Narratives: Individual interviews

We conducted semi-structured qualitative interviews (**Appendix A**) with parents who reported that their child had COVID-19. Parents were asked to share their experiences having a child with the illness. Data was analyzed thematically. Results from this study are published elsewhere (21).

### Compilation of Parents’ Narratives: Focus Groups

We conducted focus groups with parents who were recruited online via social media and through community partners to increase diversity of participants. A semi-structured interview tool guided the discussions (**Appendix B**). Data was analyzed thematically. Results from this study are published elsewhere (22).

### Prototype (Intervention) Development

Results from the qualitative interviews and focus groups were used to inform the development of infographic skeletons and video scripts. Three main topics emerged from the qualitative data, and were used as the basis for the KT tool content. These were related to 1) socialization and reintegration of children into regular activities, 2) COVID-19 vaccines for children, and 3) parenting a child who may have COVID-19. Following the completion of the infographic skeletons and scripts, researchers worked with illustrators and graphic designers to develop the tool prototypes.

### Revisions

Iterative processes were used to develop the tools whereby parents, health care professionals (HCPs), and researchers provided several rounds of feedback. Pediatric HCPs were purposively identified for their expertise on the content and were asked to provide feedback on the content and clinical accuracy of the prototypes. The research team met bi-weekly to discuss the development of the tools and to provide direction and input on all aspects of the tools. Parents from our Pediatric Parent Advisory Group (P-PAG) (19, 20) provided feedback on the content, length, stylistic elements, and any information missing from the tools.

### Usability Surveys

Parents were recruited from our Parent Advisory Groups (19, 20) via email to participate in an electronic usability survey (**Appendix I**). They were sequentially sent links over six weeks to the tool prototypes and were asked to review the tools, then complete the survey. Surveys comprised of nine, 5-point Likert items assessing: 1) usefulness, 2) aesthetics, 3) length, 4) relevance, and 5) future use. Parents were also asked to provide their positive and negative opinions of the tools via two free text boxes. Once a minimum of 30 completed surveys for each of the six tools were submitted the survey was closed.

### Data Analysis

Data from usability testing was cleaned and analyzed using SPSS v.29. Descriptive statistics and measures of central tendency were generated for demographic questions. Likert answers were given a corresponding numerical score, with 5 being “Strongly Agree” and 1 being “Strongly Disagree”, and means and standard deviations were calculated. Open-ended survey data was analyzed thematically. Comments were reviewed by the researchers and discussed to ensure changes could be made to the KT tools when possible.

### Dissemination

Once completed, the tools were disseminated via our social media channels, including Facebook(@echoKTresearch), Instagram (@echoKTofficial), and X (@echoKTresearch, @arche4evidence, @ArcheEchoKT, @cochrane_child). All tools are hosted on our website https://www.echokt.ca/tools/covid which is linked to by other trusted Canadian healthcare websites. Videos are also played across Canada through partnerships with pediatric hospitals and Alberta Health Services. We also had a targeted media campaign (TV and radio) that created further awareness of these resources.

## Results

Parents were engaged in supporting the research. A total of 27 parents were interviewed between May 2020 and April 2022. Results demonstrated that “parent experiences were diverse and multi-faceted, and their experiences evolved and shifted over the course of the pandemic.”(21). A total of 12 focus groups with 67 parents were conducted between October and December of 2021. Results demonstrated that “trying to mitigate the risk of COVID-19 infection and adhere to public health recommendations, while balancing various factors (work, online learning, and social interactions) and navigating changing information, was overwhelming for many parents” (22).

The tools (video and interactive infographic) addressed issues raised by parents regarding navigating their child’s social world, vaccination, and parenting when your child has COVID-19.

### Videos

#### 1. COVID-19 and your child’s social world

The English-language video is 3 minutes and 31 seconds in length. It includes closed captioning and outlines the story of three parents in a discussion about getting their children back into activities such as band practice and school following in-person restrictions during the pandemic. The parents discuss how their children are anxious about returning to in-person activities. One parent shared how their child was struggling with the return to school in-person and also discusses the help they received from their doctor in setting up a plan with the school to overcome challenges. Another parent discussed the symptoms of anxiety that they were seeing in their child and the ones their doctor discussed with them such as difficulty sleeping, upset stomach, being grumpier and worrying. The parent recommended talking to your health care provider if worried about your child. The narrator then discussed the differences between worrying and clinical anxiety and recommended talking to your health care provider if concerned. Screen captures of the video are included in **Appendix C**.

#### 2. COVID-19 and vaccines for children

The English-language video is 3 minutes and 41 seconds in length, and includes closed captioning. It is set at a soccer field with two parents talking about vaccinations. The parent of Remi chose to have her vaccinated while Charlie’s parent was unsure about vaccinating Charlie. In the conversation between the two parents, Charlie’s dad asked several questions about the vaccine and Remi’s parent provided evidence-based answers. Remi’s parent discussed how she went to her doctor to discuss the vaccine. Charlie’s father stated that he felt COVID-19 was like a cold and talked about how Charlie hates needles. Remi’s mother then discussed long COVID and needle pain strategies. The video presents credible websites that offer evidence-based information to answer questions about vaccines, their development, and approval processes. Screen captures of the video are included in **Appendix D**.

### 3. COVID-19 and parenting a child who may have COVID

The English-language video is 3 minutes and 52 seconds in length, and includes closed captioning. It is set at an outdoor swimming pool, where 3 parents are watching their children in swimming lessons. A couple is talking to a parent about their experiences of both their children having had COVID-19. They discuss the symptoms their children had, and that they contacted their doctor’s office for advice from the nurse, including how to manage fevers and which symptoms would indicate the need to go to the emergency department. The parents shared how their children felt emotionally during the time they had COVID, and how confusing and frustrating it was to find inconsistent information on isolation. The other parent discussed how isolating in her apartment would be a challenge, and how not being able to go to work would be financially challenging. The parents discussed the challenges of getting their children back to normal activities after recovery. The father stressed the need to stay home if sick, keep children home if they are sick even though it can be challenging, and to reach out for help if their children do get COVID-19. Screen captures of the video are included in **Appendix E**.

### Infographics

Three interactive infographics were developed in the same format as other infographics in our suite of tools (https://www.echokt.ca/tools/). This style is unique to our research program and was developed and evaluated over the course of several years. (20) The infographics look similar to webpages and allow users to scroll through the information at their own pace and select the information that is relevant to their situation. The topics presented in the infographics mirror those in the videos but the infographics provide more details and recommendations about where to find additional reliable information. The ability for parents and caregivers to control what they view based on their informational needs differentiates the infographics from the videos.

#### 1. COVID-19 and your child’s social world

This interactive infographic is comprised of five major sections: (1) questions about school (2) colds, flus and other illnesses (3) extracurricular activities (4) concerns about mental health, and (5) useful links. The sections begin with a quote from a story shared by parents in both audio and visual formats. Included in the sections are additional questions that parents may want to ask regarding their child’s activities. The section on colds, flus and other illnesses includes a section on how these illnesses may affect respiratory and gastrointestinal systems. Each of the sections has links to relevant information. The ending includes links to references and additional resources. Screen captures are included in **Appendix F**.

#### 2. COVID-19 and vaccines for children

This interactive infographic mirrors the information provided in the video and is comprised of five major sections: (1) vaccine development (2) talking to your child (3) supporting your child (4) trustworthy information and (5) useful links. The sections begin with a recorded quote from a story shared by the parents that has both an audio and a visual component. The section on vaccine development goes through the four stages of how vaccines are developed and approved in Canada. The section on supporting your child contains tips for managing vaccine stress and needle pain in children. The section on trustworthy information includes definitions of misinformation and disinformation, along with links to trusted resources. The useful links section at the end includes the references cited in the infographic and links to additional resources. Screen captures are included in **Appendix G**.

#### 3. COVID-19 and parenting a child who may have COVID-19

This interactive infographic mirrors the information provided in the video and is comprised of six major sections: (1) having COVID-19 (2) symptoms (3) what to do (4) managing stress (5) getting help and (6) resources. Most of the sections begin with a recorded quote from a story shared by a parent that has both an audio and a visual component. The section on having COVID-19 includes links to government websites that provide up-to-date information, as the information changes frequently. The section on symptoms relayed information about common symptoms of COVID-19 in children. The section on what to do includes the list of symptoms that would necessitate taking a child to the emergency department. The resource section includes references and links to additional resources. Screen captures are included in **Appendix H**.

### Usability Testing Results

Parents were engaged in usability testing and were quick to participate through the completion of surveys. We received at least 30 responses for each KT tool. All 6 KT tools were rated highly, with averages in mean scores ranging from 3.42 to 4.48 out of 5.

The tools about COVID-19 and your child’s social world were evaluated by 63 parents (**Table 1**). Mean scores across the usability items were 3.59 to 4.38 for the infographic, and 3.42 to 4.42 for the video (**Table 2**). The tools about COVID-19 vaccines for children were evaluated by 62 parents (**Table 3**). Mean scores across the usability items were 3.97 to 4.48 for the infographic, and 3.65 to 4.31 for the video (**Table 4**). The tool about parenting a child with COVID-19 were evaluated by 60 parents (**Table 5**). Mean scores across the usability items were from 3.87 to 4.40 for the infographic, and 3.47 to 4.47 for the video (**Table 6**). Comments from the two open-text questions at the end of the usability survey were constructive and mainly positive.

**Table 1.** Demographic characteristics of parents who assessed the usability of the digital tools about COVID-19 and your child’s social world (n=63).

| Characteristic | n (%) |
| --- | --- |
| Gender |  |
| Female | 49 (78) |
| Male | 14 (12) |
| Ethnicity |  |
| East Asian | 7 (11) |
| Indigenous | 0 (0) |
| South Asian | 6 (10) |
| White/Caucasian | 45 (72) |
| Not Listed or Other | 3 (4) |
| Prefer not to answer | 2 (3) |
| Age |  |
| Less than 20 | 0 (0) |
| 20-30 years | 6 (10) |
| 31-40 years | 31 (49) |
| 41-50 years | 16 (25) |
| 51-60 years | 10 (16) |
| Supportive adult in everyday life |  |
| Yes | 58 (92) |
| No | 5 (8) |
| Education |  |
| Some high school | 0 (0) |
| High school diploma | 1 (2) |
| Some post-secondary | 4 (6) |
| Post-secondary certificate/diploma | 5 (8) |
| Post-secondary degree | 31 (49) |
| Graduate degree | 21 (33) |
| Prefer not to answer | 1 (2) |
| Household Income |  |
| Less than \$25,000 | 0 (0) |
| \$25,000-\$49,000 | 4 (6) |
| \$50,000-\$74,000 | 4 (6) |
| \$75,000-\$99,000 | 16 (25) |
| \$100,000-\$149,000 | 16 (25) |
| \$150,000 and over | 14 (12) |
| Prefer not to answer | 9 (14) |
| Geographical Area |  |
| Inner City | 28 (44) |
| Suburb | 28 (44) |
| Town | 6 (10) |
| Farm/rural | 1 (2) |
| Number of Children in the Family |  |
| 1 | 23 (36) |
| 2 | 28 (44) |
| 3 | 7 (11) |
| 4 | 2 (3) |
| 5+ | 2 (3) |
| Missing | 1 (2) |
| Primary language(s) spoken in the home |  |
| English | 53 (84) |
| English and French | 2 (3) |
| English and Mandarin | 2 (3) |
| Mandarin | 2 (3) |
| Nepali | 4 (6) |
| Were you born in Canada? |  |
| Yes | 51 (81) |
| No | 12 (19) |

**Table 2.** Means (SD) of participant responses to the usability survey of the digital tools about COVID-19 and your child’s social world.

| Usability Measures | Infographic<br>(n=32) | Video<br>(n=31) |
| --- | --- | --- |
| It is useful | 4.25 (.72) | 4.29 (.69) |
| It provides information that is relevant to me as a parent | 4.25 (.67) | 4.23 (.80) |
| It is simple to use | 4.09 (.93) | 4.35 (.49) |
| I can use it without written instructions or additional help | 4.38 (.70) | 4.42 (.56) |
| Its length is appropriate | 3.59 (1.16) | 4.03 (1.16) |
| It is aesthetically pleasing (i.e. images, colours, etc.) | 4.06 (.98) | 3.90 (.94) |
| It helps me make decisions about my child's health | 4.13 (.87) | 3.77 (.84) |
| I would use it in the future | 3.77 (1.12) | 3.42 (1.06) |
| I would recommend it to a friend | 3.91 (1.09) | 3.83 (1.12) |

**Table 3.** Demographic characteristics of parents who assessed the usability of the digital tools about COVID-19 and vaccines for children (n=62).

| Characteristic | n (%) |
| --- | --- |
| Gender |  |
| Female | 43 (69) |
| Male | 19 (31) |
| Ethnicity |  |
| Black | 2 (3) |
| East Asian | 7 (11) |
| Indigenous | 0 (0) |
| Middle Eastern | 2 (3) |
| South Asian | 6 (10) |
| White/Caucasian | 40 (65) |
| Not Listed or Other | 3 (5) |
| Prefer not to answer | 2 (3) |
| Age |  |
| Less than 20 | 0 (0) |
| 20-30 years | 6 (10) |
| 31-40 years | 28 (45) |
| 41-50 years | 20 (32) |
| 51 and older | 8 (13) |
| Did not answer | 1 (2) |
| Supportive adult in everyday life |  |
| Yes | 58 (94) |
| No | 4 (6) |
| Education |  |
| Some high school | 0 (0) |
| High school diploma | 0 (0) |
| Some post-secondary | 2 (3) |
| Post-secondary certificate/diploma | 7 (11) |
| Post-secondary degree | 30 (48) |
| Graduate degree | 20 (32) |
| Prefer not to answer | 2 (3) |
| Household Income |  |
| Less than \$25,000 | 0 (0) |
| \$25,000-\$49,000 | 3 (5) |
| \$50,000-\$74,000 | 4 (6) |
| \$75,000-\$99,000 | 6 (10) |
| \$100,000-\$149,000 | 23 (37) |
| \$150,000 and over | 13 ((21) |
| Prefer not to answer | 11 (18) |
| Geographical Area |  |
| Inner City | 25 (40) |
| Suburb | 27 (44) |
| Town | 7 (11) |
| Farm/rural | 1 (2) |
| Other | 2 (3) |
| Number of Children in the Family |  |
| 1 | 18 (29) |
| 2 | 16 (26) |
| 3 | 2 (3) |
| 4 | 4 (6) |
| 5+ | 9 (15) |
| Missing response | 2 (3) |
| Primary language(s) spoken in the home |  |
| English | 56 (90) |
| English and Mandarin | 2 (3) |
| English and Nepali | 1 (2) |
| Nepali | 2 (3) |
| Were you born in Canada? |  |
| Yes | 49 (79) |
| No | 13 (21) |

**Table 4.**
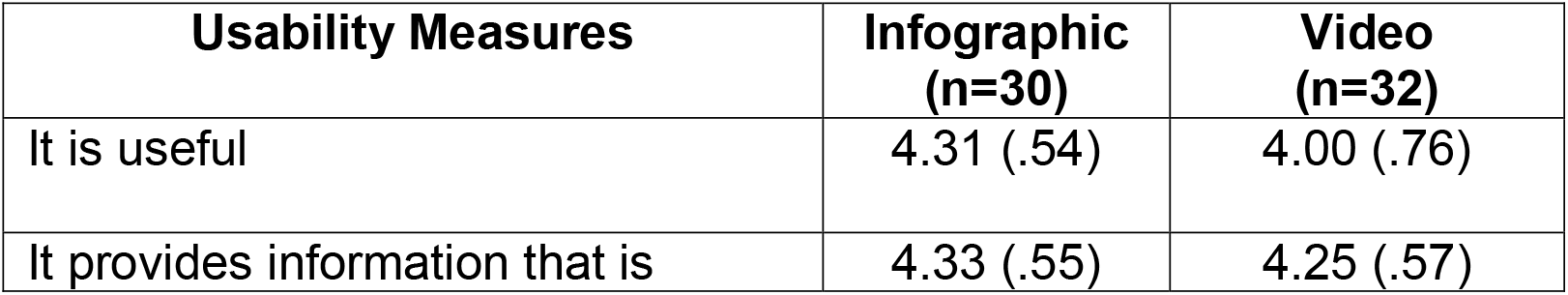
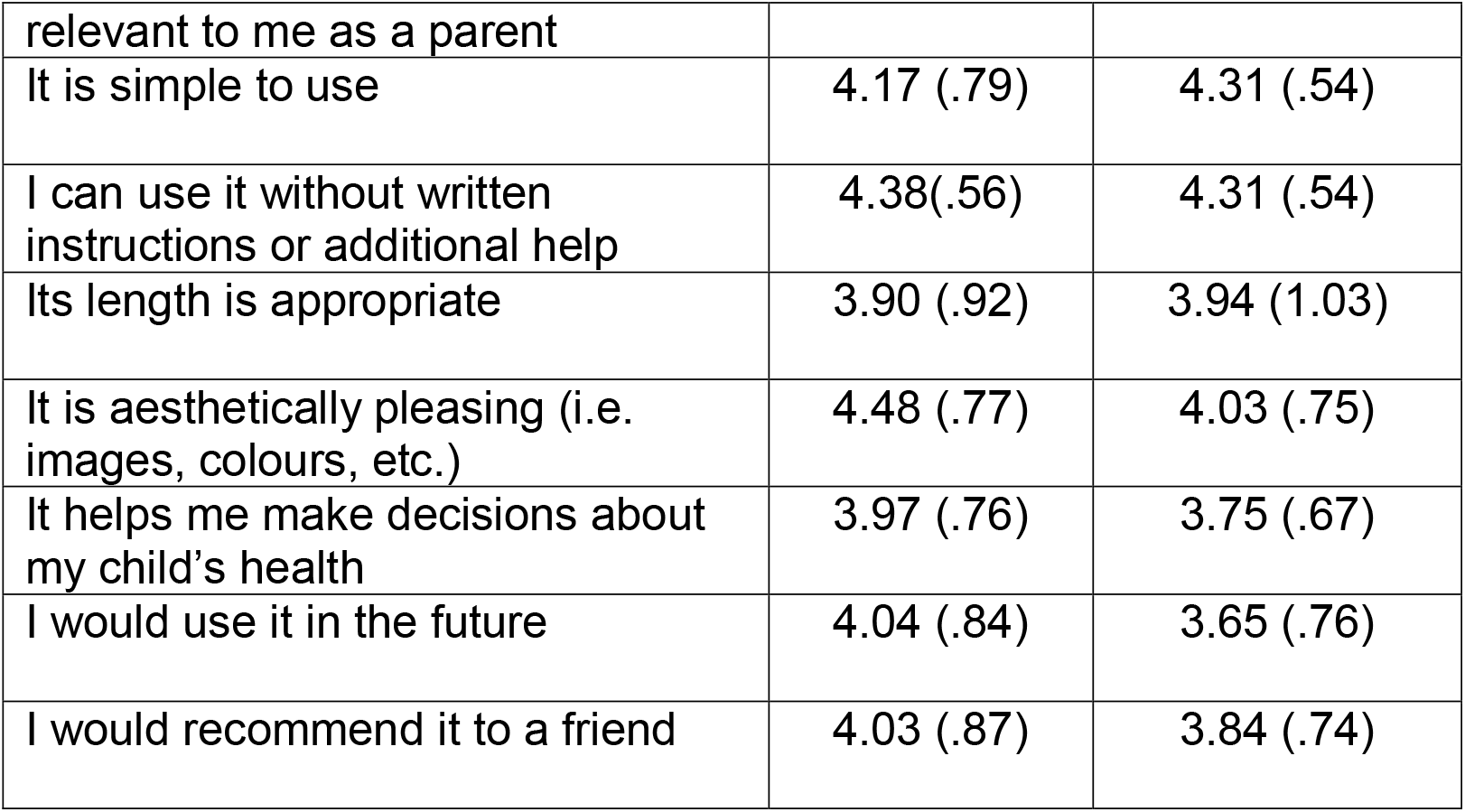
Means (SD) of participant responses to the usability survey of the digital tools about COVID-19 and vaccines in children.

**Table 5.** Demographic characteristics of parents who assessed the usability of the digital tools about parenting a child who may have COVID-19 (n=60).

| Characteristic | n (%) |
| --- | --- |
| Gender |  |
| Female | 46 (77) |
| Male | 13 (22) |
| Non-binary | 1 (2) |
| Would you describe yourself as transgender? |  |
| Yes | 1 (2) |
| No | 58 (96) |
| Blank | 1 (2) |
| Ethnicity |  |
| Black | 1 (2) |
| East Asian | 11 (18) |
| Indigenous | 0 (0) |
| Middle Eastern | 0 (0) |
| South Asian | 7 (12) |
| White/Caucasian | 40 (67) |
| Not Listed or Other | 0 (0) |
| Prefer not to answer | 1(2) |
| Age |  |
| Less than 20 | 0 (0) |
| 20-30 years | 4 (7) |
| 31-40 years | 20 (33) |
| 41-50 years | 30 (50) |
| 51 and older | 6 (10) |
| Did not answer | 0 (0) |
| Supportive adult in everyday life |  |
| Yes | 55 (92) |
| No | 4 (7) |
| Prefer not to answer | 1 (2) |
| Education |  |
| Some high school | 0 (0) |
| High school diploma | 0 (0) |
| Some post-secondary | 4 (7) |
| Post-secondary certificate/diploma | 7 (12) |
| Post-secondary degree | 25 (42) |
| Graduate degree | 23 (38) |
| Prefer not to answer | 1 (2) |
| Household Income |  |
| Less than \$25,000 | 0 (0) |
| \$25,000-\$49,000 | 4 (7) |
| \$50,000-\$74,000 | 3 (5) |
| \$75,000-\$99,000 | 10 (17) |
| \$100,000-\$149,000 | 25 (42) |
| \$150,000 and over | 10 (17) |
| Prefer not to answer | 8 (13) |
| Geographical Area |  |
| Inner City | 33 (55) |
| Suburb | 25 (42) |
| Town | 2 (3) |
| Farm/rural | 0 (0) |
| Other | 0 (0) |
| Number of Children in the Family |  |
| 1 | 12 (20) |
| 2 | 32 (53) |
| 3 | 7 (12) |
| 4 | 7 (12) |
| 5+ | 2 (3) |
| Primary language(s) spoken in the home |  |
| English | 52 (87) |
| English and Mandarin | 2 (3) |
| Nepali | 4 (7) |
| Mandarin | 1 (2) |
| Were you born in Canada? |  |
| Yes | 50 (83) |
| No | 9 (15) |
| Prefer not to answer | 1 (2) |

**Table 6.**
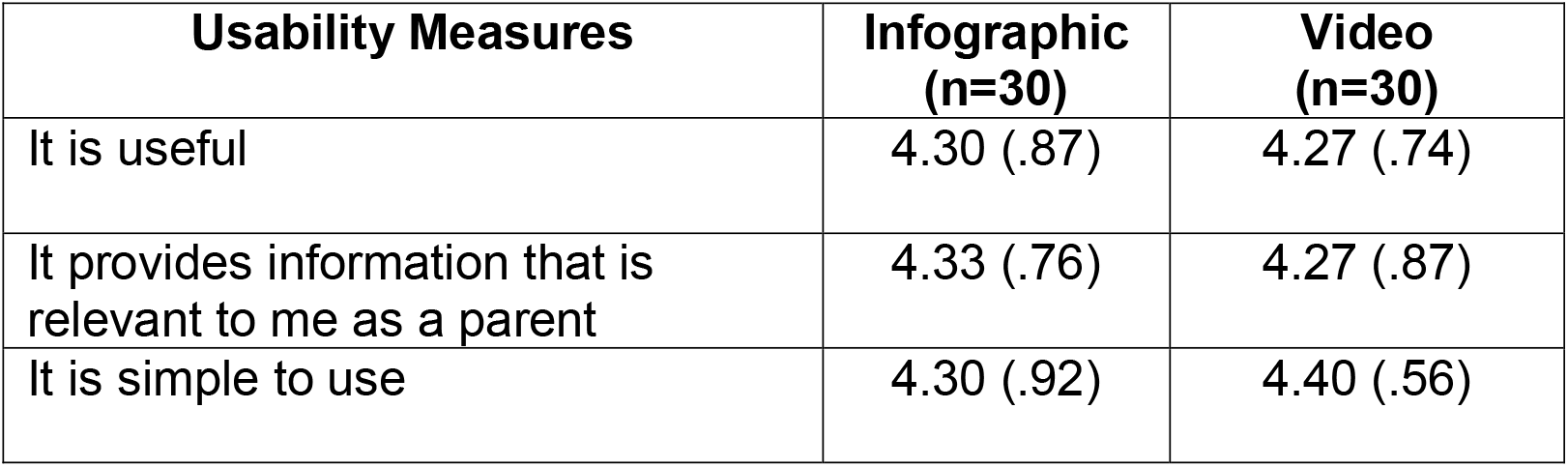
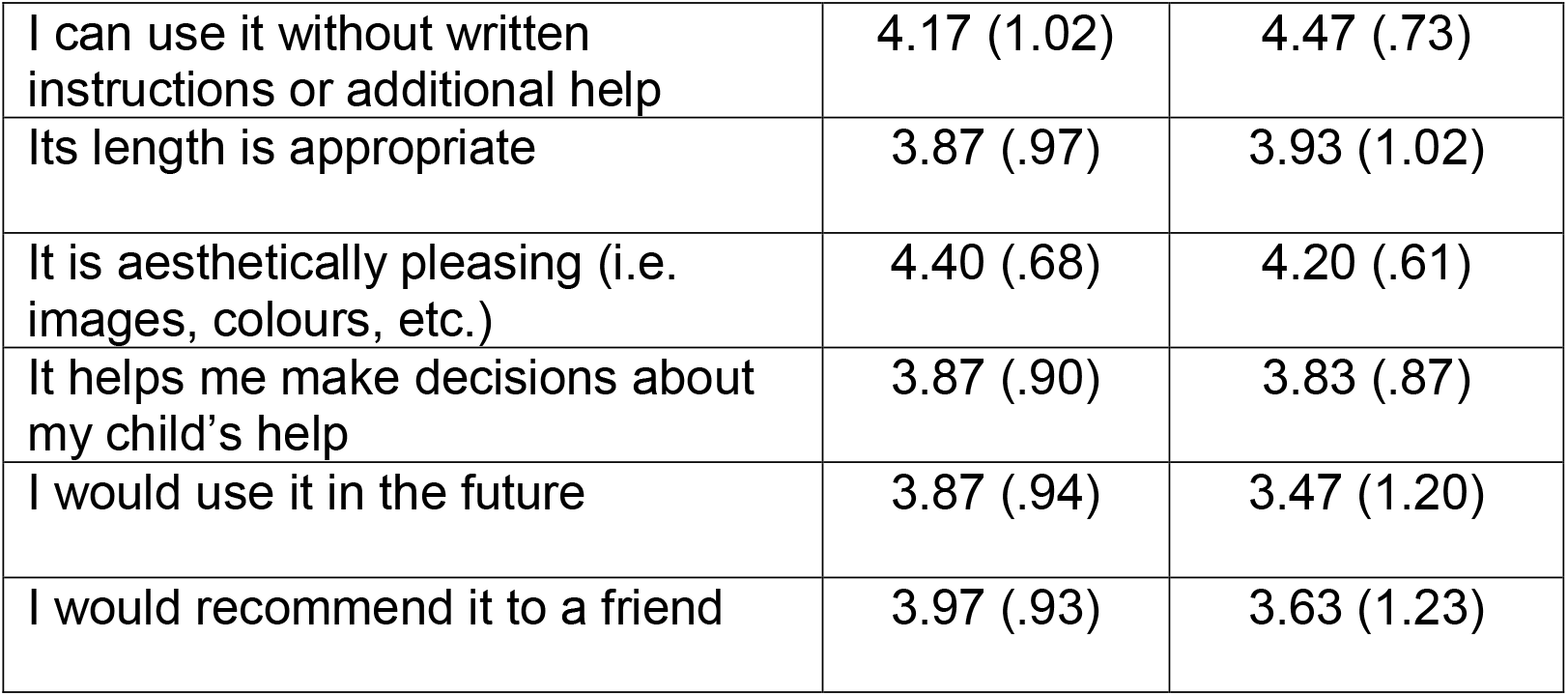
Means (SD) of participant responses to the usability survey of the digital tools about parenting a child who may have COVID-19 (n=60).

| Usability Measures | Infographic<br>(n=30) | Video<br>(n=30) |
| --- | --- | --- |
| It is useful | 4.30 (.87) | 4.27 (.74) |
| It provides information that is relevant to me as a parent | 4.33 (.76) | 4.27 (.87) |
| It is simple to use | 4.30 (.92) | 4.40 (.56) |
| I can use it without written instructions or additional help | 4.17 (1.02) | 4.47 (.73) |
| Its length is appropriate | 3.87 (.97) | 3.93 (1.02) |
| It is aesthetically pleasing (i.e. images, colours, etc.) | 4.40 (.68) | 4.20 (.61) |
| It helps me make decisions about my child's help | 3.87 (.90) | 3.83 (.87) |
| I would use it in the future | 3.87 (.94) | 3.47 (1.20) |
| I would recommend it to a friend | 3.97 (.93) | 3.63 (1.23) |

Following refinement based on usability testing results, the KT tools were made publicly available in December 2022 on our website (echokt.ca) and shared with relevant stakeholders.

## Conclusion

Working together with parents, we developed, evaluated and revised six KT tools to help parents navigate their information needs and concerns related to COVID-19 and the pandemic. We conducted focus groups with interested parents about their experiences and information needs related to the pandemic with specific questions about public health measures (e.g., social distancing, vaccinations). In addition, we conducted individual interviews with parents whose children had been diagnosed with COVID-19 to understand their experiences and information needs related to having a child diagnosed with COVID-19.

We integrated parents’ experiences and information needs with best available evidence to develop six KT tools: one video and one interactive infographic on each of three topics, parenting a child who may have covid-19, navigating a child’s social world during the pandemic and beyond, and covid-19 and vaccines for children. By working with clinical experts in child health, we ensured the tools were clinically accurate and reflected best evidence. The six tools were evaluated for usability by parents from our existing networks, and results suggest that parents found the tools to be highly useful in sharing complex information about COVID-19.

Results from the development and evaluation of these novel KT tools suggests that fostering patient engagement throughout the development of KT tools is a successful model for the development of highly usable and acceptable resources.

## Data Availability

All data produced in the present study are available upon reasonable request to the authors

## Author Contributions

This research was conducted under the supervision of Drs. Shannon D. Scott (SDS) and Lisa Hartling (LH), PIs for **translation Evidence in Child Health to enhance Outcomes** (ECHO) Research and the **Alberta Research Centre for Health Evidence** (ARCHE), respectively. Both PIs designed the research and obtained research funding through the Canadian Institutes of Health Research (CIHR).

SDS and LH designed and supervised all aspects of tool development and evaluation. Sarah Elliott (SE) was involved in design of the project, oversaw the focus group component and development of tools, and supervised research staff. Lisa Knisley (LK) was involved in design of the project and evaluation of tools.

SDS and Kathy Reid (KR) conducted and analyzed qualitative interviews with parents whose children had COVID-19. SE conducted and analyzed qualitative focus group discussions with parents.

KR conducted usability testing of all tools with parents from our Pediatric Parent Advisory Group (P-PAG) and our Pediatric Parent Consultation Network (P-PCN). KR analyzed usability data for all tools.

All authors contributed to the writing of this report and provided substantial feedback.

This work was funded by:

**The Canadian Institutes of Health Research:**

- Scott, S.D., Hartling, L., Elliott, S.A. & Knisley, L (2021) Overcoming emergency COVID-19 challenges with relatable science: developing and evaluating knowledge tools for Canadians ($272, 470). July 2021 – March 2024.

AND

- Scott, S.D. (co-PI) & Hartling, L. (co-PI), Ali, S., Currie, G., Dyson, M., Fernandes, R., Fleck, B., Freedman, S., Jabbour, M., Johnson, D., Junker, A., Klassen, T., Maynard, D., Newton, A., Plint, A., Richer, L., Robinson, J., Robson, K., Vandall-Walker, V. [all collaborators listed in alphabetical order]. (2016) Integrating evidence and parent engagement to optimize children’s healthcare. CIHR Foundation Scheme ($2,500,000). July 2016 – March 2027.
- Hartling, L. & Scott, S.D (2018). Distinguished Research Funding. Stollery Children’s Hospital Foundation and the Women and Children’s Health Research Institute ($1,000,000), September 2018 – January 2025.

**This report should be cited as:**

Scott, S.D., Elliott, S.A., Reid, K., Knisley, L., Hartling, L. (2024). The development and usability testing of six arts-based knowledge translation tools for parents about COVID-19. Internal Technical Report. ECHO Research, University of Alberta.

## Available at

*http://www.echokt.ca/research/technical-reports/*

## Acknowledgements

We would like to thank Kelsey Wright for their support on this project, particularly during focus group data collection and analysis. We would also like to thank the many parents who shared their experiences and time with us.

## Other Outputs from this Project and Related Works

## Appendix A: Parent Experiences Qualitative Interview Guide

### Understanding the Information Needs & Experiences of Parents with a child with COVID19. Semi-Structured Interview Guide

1. Tell me about <u>your experience</u> having your child diagnosed with COVID19
  a. Probe for:
    i. the events <u>leading up</u> to your child being diagnosed. (Do they know how/when they became infected)
    ii. signs/symptoms your child was experiencing (Child’s health)
      1. Child’s age
    iii. visits to healthcare professionals or telehealth to understand what was happening. (Probe number of visits/telehealth, types of healthcare settings they visited, what did they do? Medications, assessments; were other diagnoses given to your child?)
    iv. things you tried at home to decrease the symptoms (did you ask family members, friends, public health, grandparents for help to understand what could be wrong with the child).
    v. Did you look for information at this time? (where did you look for information)
    vi. how did <u>your child feel</u> during this time? (emotions, mental state)
    vii. how did <u>you feel</u> during this time? (emotions, mental state)
2. Tell me <u>when your child was diagnosed</u> with COVID.
  a. Probe for:
    i. who diagnosed your child with COVID?
    ii. did your child go to the Emergency Department? Where were they diagnosed
    iii. how did they diagnosis your child with COVID19 (what tests did they do? How long did you have to wait to receive the test results? How did you feel while you were waiting?)
3. Tell me what <u>happened after</u> they were diagnosed with COVID19?
  a. Probe for:
    i. What medical care did you your child receive (hospitalized? Went to ED, went to family physician, etc)?
    ii. Sense of child’s trajectory of illness (resp status, energy level, difficulty breathing, appetite, other symptoms)
    iii. medications ordered? (tell me about the medications and response to medications)
    iv. education given for day-to-day management
    v. changes that the family needed to make for the child with COVID19.
4. Tell me about the <u>when your child was able to go home.</u>
  a. When did your child get discharged home? How did you feel about managing your child’s health needs – Probe for parent’s level of comfort/knowledge/skill being able to manage their child’s needs.
5. Tell me about the rest of your family and how they were affected with your child’s diagnosis of COVID19.
  a. Probe for:
    i. Did the family need to make significant lifestyle or living arrangement changes? (quarantine/self isolation; how did you and your family manage this; how was it having parents and children in the home together for so long).
    ii. Did other family members get COVID19? How did you protect others from getting sick?
    iii. How did the diagnosis affect parent/caregiver work?
    iv. How did other family members (especially other children) react to the diagnosis? What challenges were associated with this?
6. How did your child adapt to having COVID?
  a. Probe for:
    i. Isolation at home/hospital
    ii. Mental/psychology impact of having COVID19
    iii. Not seeing friends/family members
    iv. No school/sports/other outside interests
7. Do you feel that you had <u>adequate knowledge</u> about COVID19 to understand what was happening to your child and to manage their illness?
8. Where did you <u>look for COVID19 information</u>?
  a. Probe for what their preferred sources and formats of health information are.
9. Were all of your <u>questions/info</u> needs about COVID19 <u>answered</u>?
10. Was there <u>information</u> that you wished you had about COVID19 but <u>could not find</u>? Tell me about that.

## Appendix B: Focus Group Qualitative Interview Guide

### Understanding parents’ experiences and information needs during the COVID19 pandemic. Semi Structured Focus Group Discussion Guide

1. Tell me about your experiences as a parent during the COVID19 pandemic.
  a. What were your concerns at the beginning of the pandemic?
  b. How did your experiences change throughout the pandemic?
  c. Did anyone in your family have COVID?
2. Tell me about your child’s experiences through the pandemic.
  a. What were your child’s concerns during this time?
  b. How did you address your child’s concerns?
3. What are your current concerns about COVID19?
  a. Do you have any concerns about your children being back in school or daycare?
  b. What are your experiences with public health restrictions in extra curricular activities for children?
  c. Do you have any concerns about the long-term effects of COVID?
4. What are your thoughts about the public health measures to prevent the spread of COVID19?
  a. What are your opinions on social distancing?
  b. What are your opinions of masks mandates?
  c. Are there any other measures or restrictions you have encountered?
5. How do your perspectives of public health measures align with your friends, family, and community members?
  a. How do you handle conflicting perspectives in your social circle?
6. Where do you seek information about the COVID19 virus?
  a. How did you evaluate the accuracy of the information?
  b. Did you experience any conflicting information about the COVID19 virus?
7. What supports and information would be helpful for you in your decisions related to COVID19?

## Appendix C: Video Screenshots – COVID-19 and your child’s social world

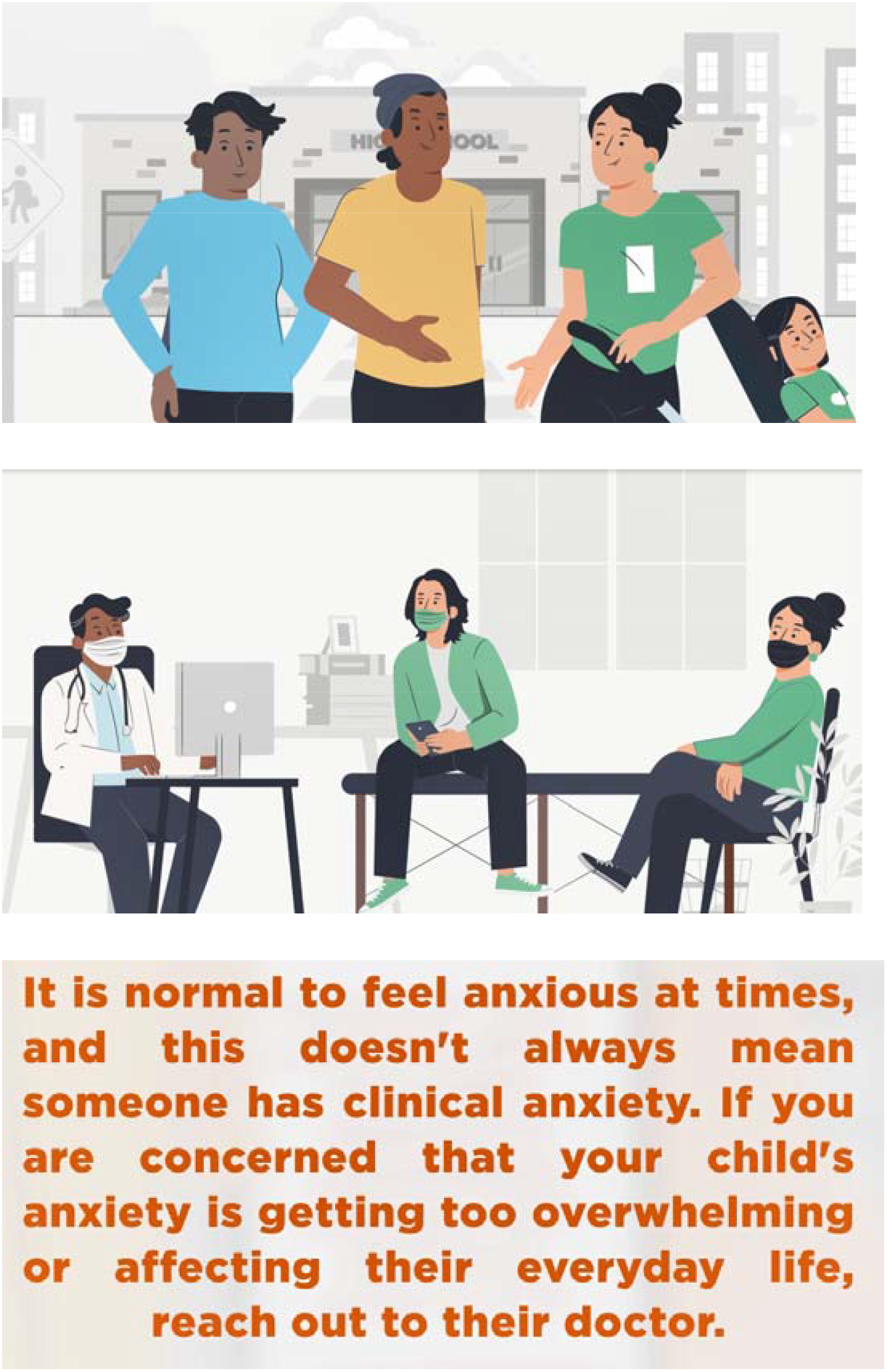

## Appendix D: Video Screenshots – COVID-19 and vaccines for children

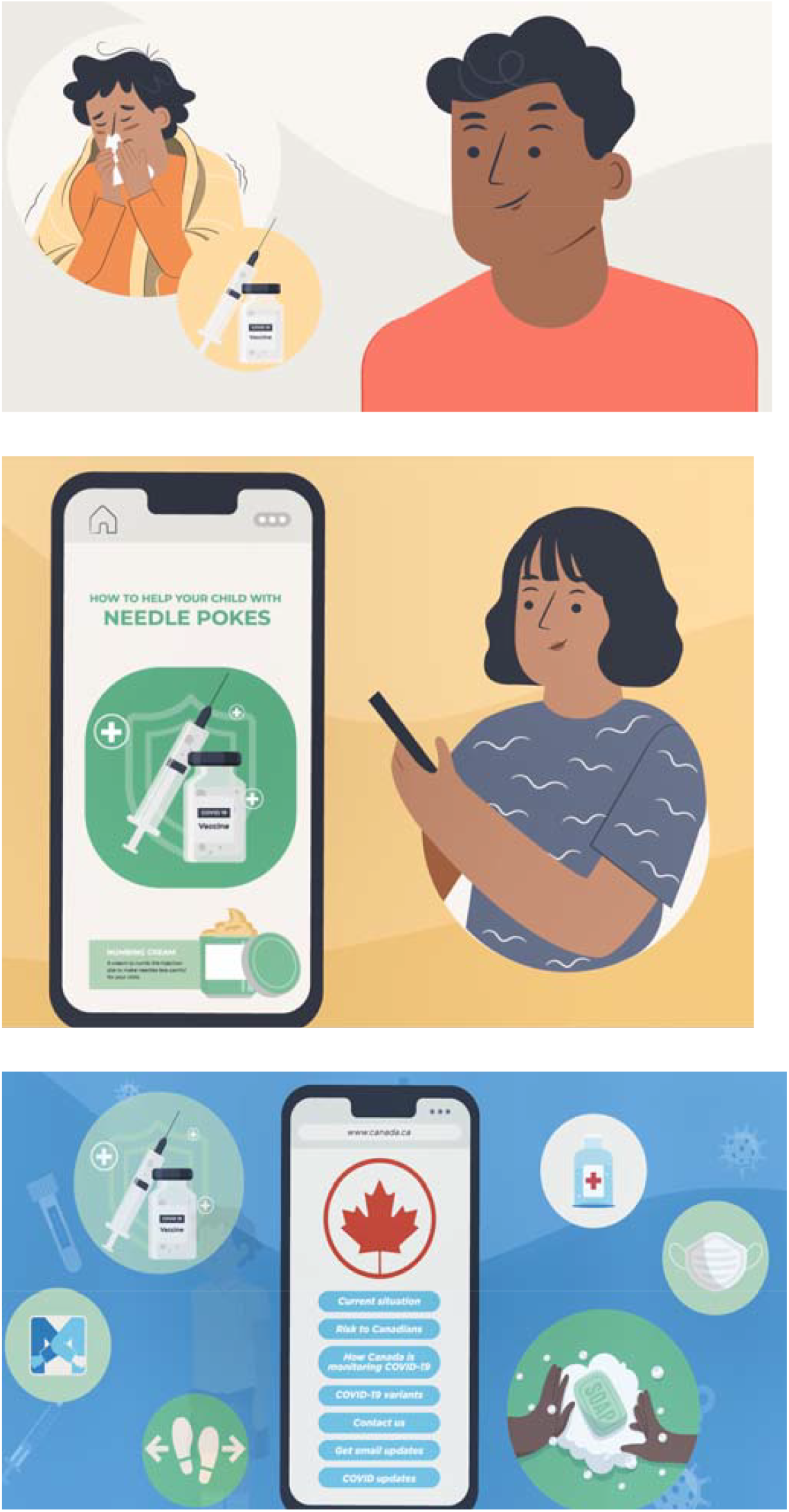

## Appendix E: Video Screenshots – COVID-19 and parenting a child who may have COVID

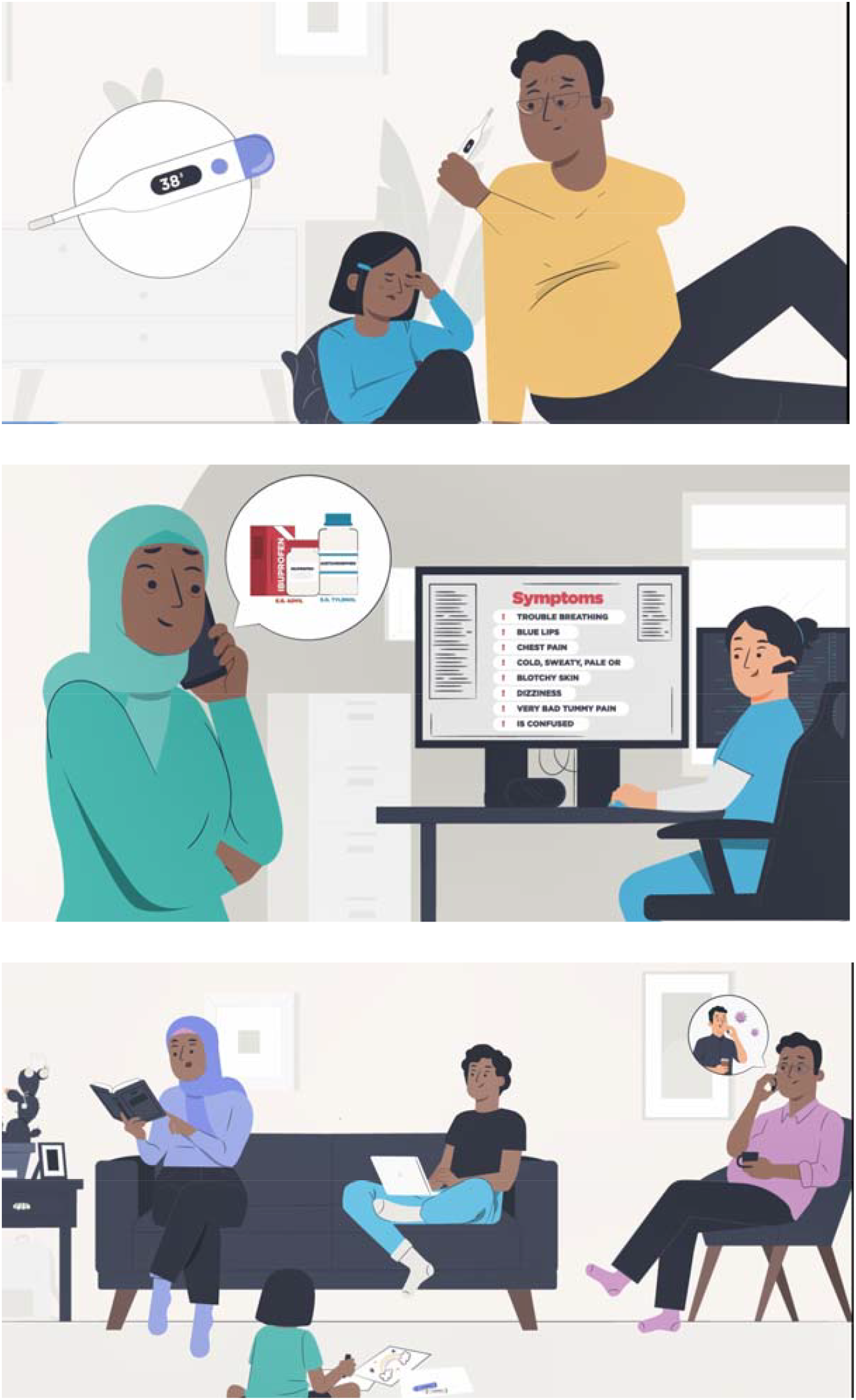

## Appendix F: Infographic Screenshots – COVID-19 and your child’s social world

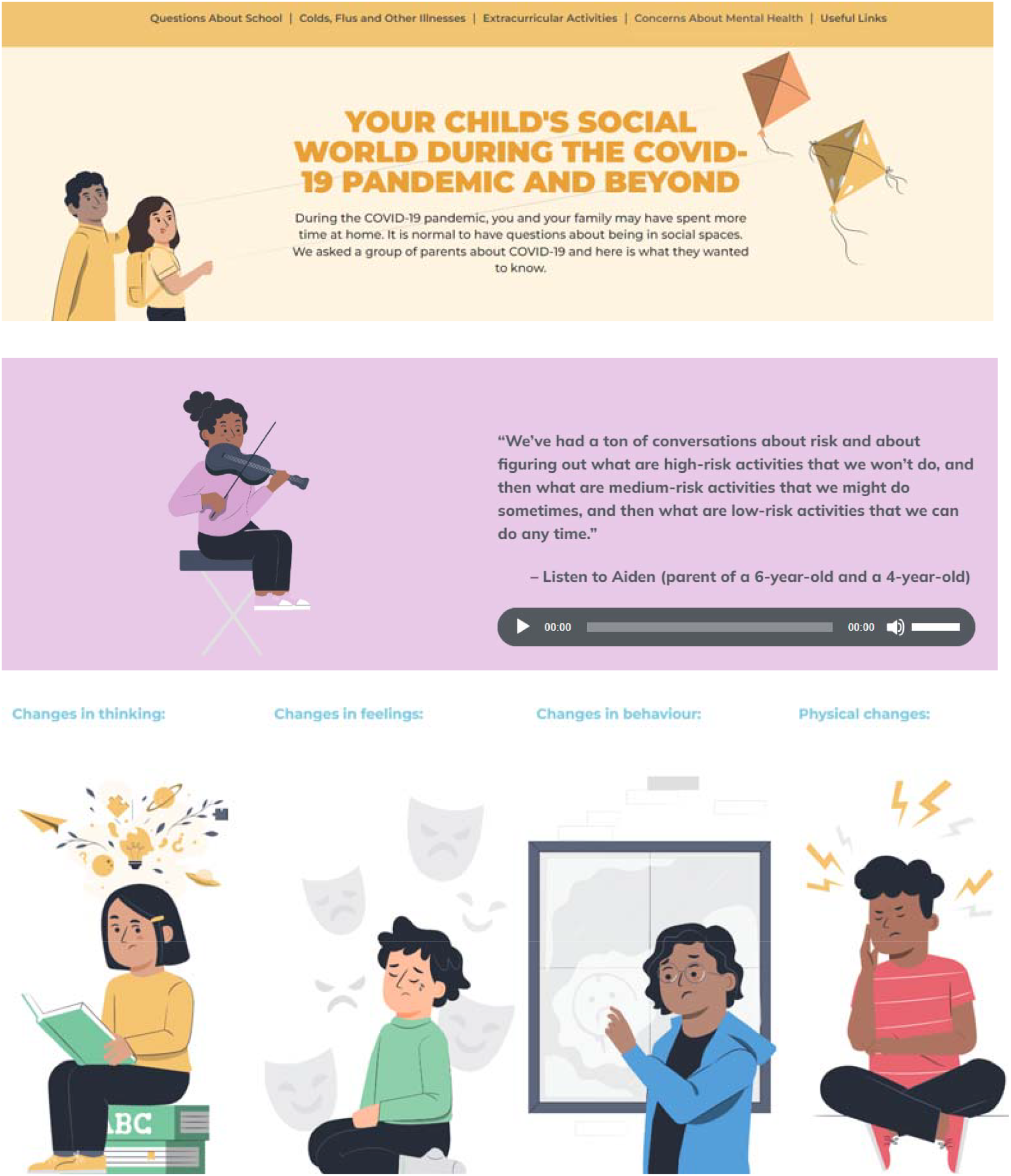

## Appendix G: Infographic Screenshots – COVID-19 and vaccines for children

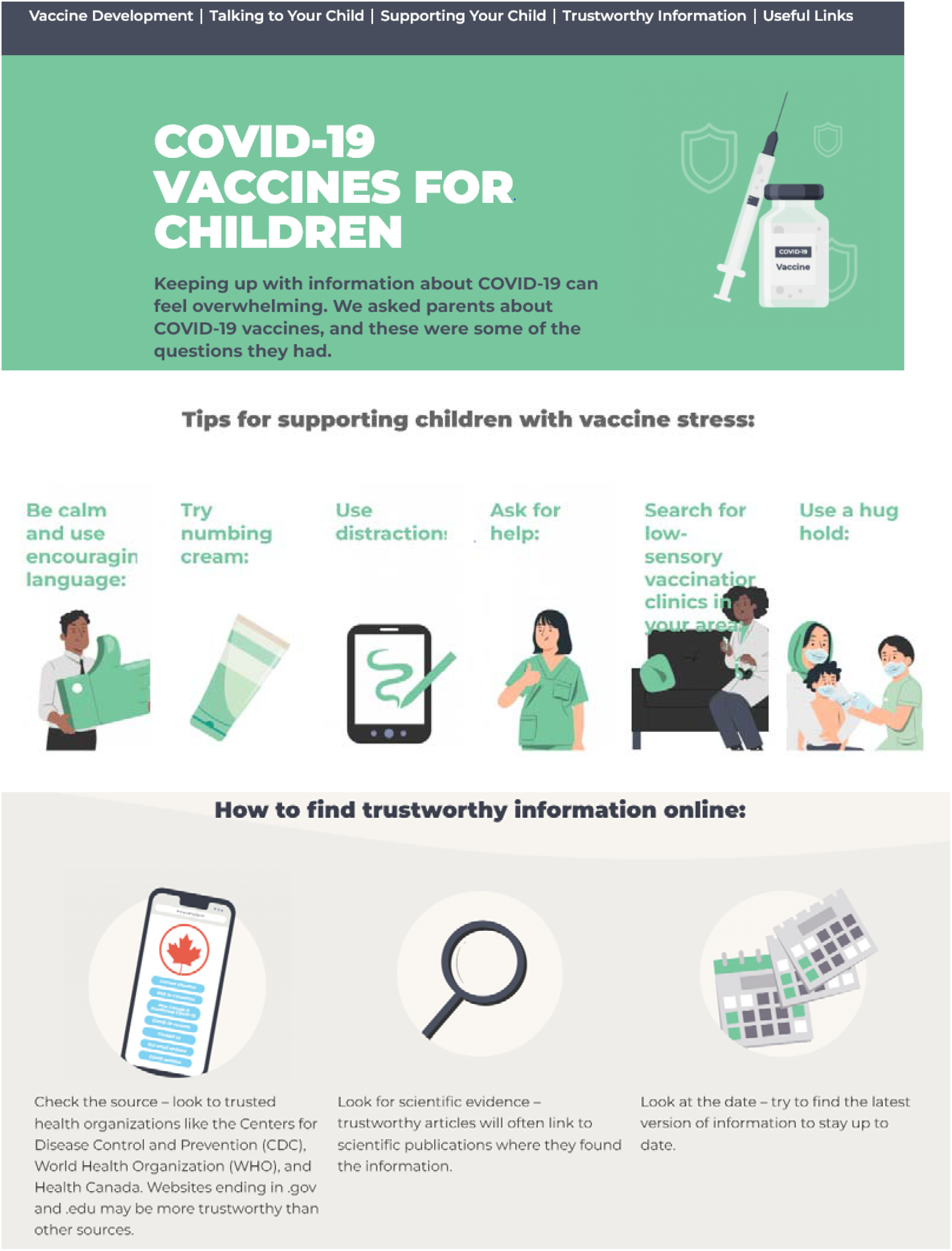

## Appendix H: Infographic Screenshots – COVID-19 and parenting a child who may have COVID

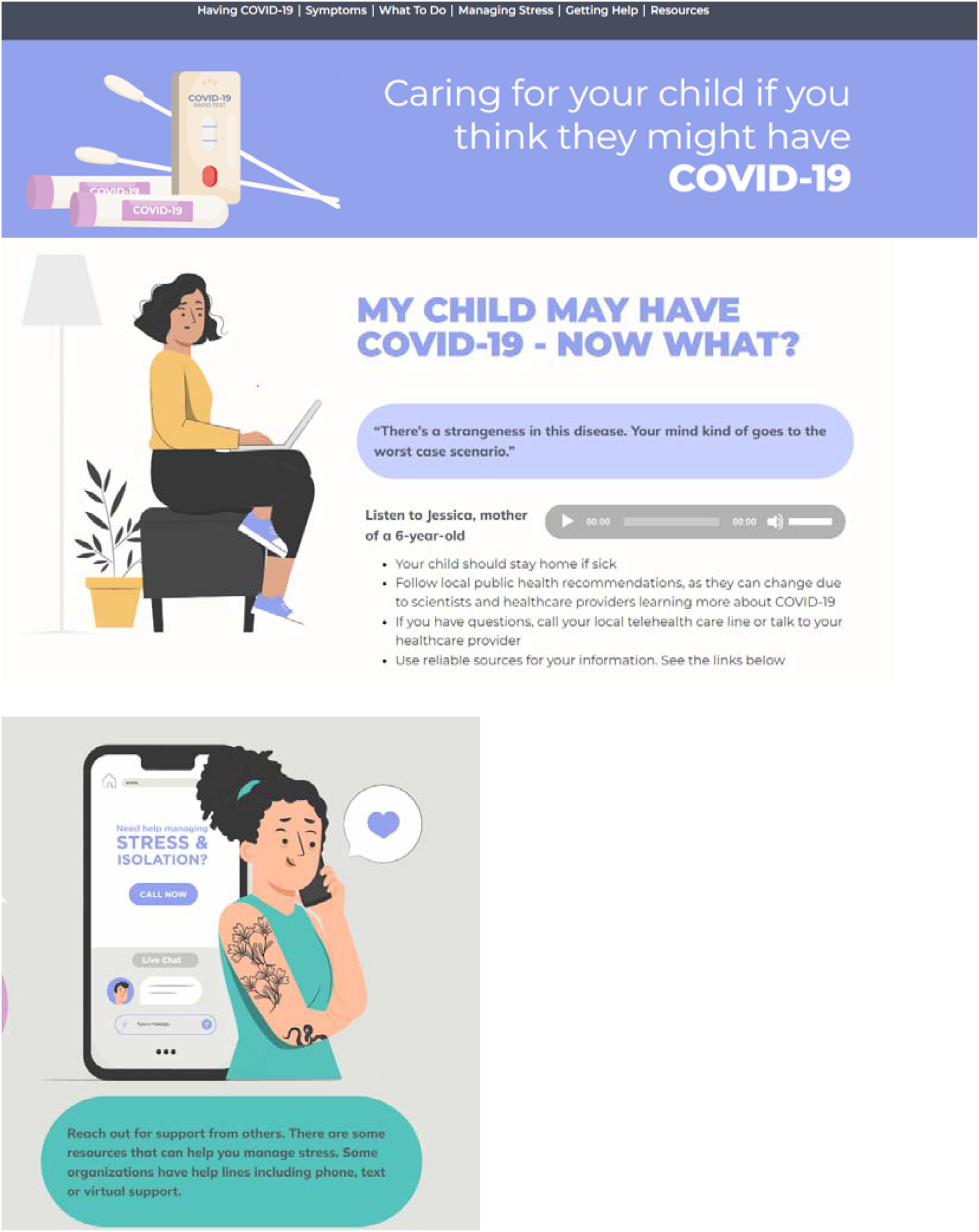

## Appendix I: Usability Survey

SECTION 1: Demographics

1. a. Which gender do you identify with most?
  ◻ Male
  ◻ Female
  ◻ Non-binary
  ◻ Two-spirit
  ◻ Other:
  ◻ Prefer not to answer
  1) b. Would you describe yourself as transgender?
  ◻ Yes
  ◻ No
  ◻ Prefer not to answer
2. Which ethnicities best describe you? *Please select all that apply*.
  ◻ Asian
  ◻ African American or African Canadian
  ◻ Black
  ◻ First Nations
  ◻ Hispanic or Latino
  ◻ Métis
  ◻ Middle Eastern or North African
  ◻ South Asian
  ◻ Southeast Asian
  ◻ White or Caucasian
  ◻ Not listed:
  ◻ Prefer not to answer
3. What is your Age?
  ◻ Less than 20 years old
  ◻ 20-30 years
  ◻ 31-40 years
  ◻ 41-50 years
  ◻ 51 years and older
4. What is your Marital Status?
  ◻ Married/Partnered
  ◻ Single
5. What is your gross annual household income?
  ◻ Less than $25,000
  ◻ $25,000-$49,999
  ◻ $50,000-$74,999
  ◻ $75,000-$99,999
  ◻ $100,000-$149,999
  ◻ $150,000 and over
  ◻ Prefer not to answer
6. What is your highest level of education?
  ◻ Some high school
  ◻ High school diploma
  ◻ Some post-secondary
  ◻ Post-secondary certificate/diploma
  ◻ Post-secondary degree
  ◻ Graduate degree
  ◻ Other:
7. Where does your household live
  ◻ City
  ◻ Suburb
  ◻ Town
  ◻ Farm
  ◻ Other:
8. What is your relationship to the child that you have brought to the emergency department?
  ◻ Parent
  ◻ Grandparent
  ◻ Other family member
  ◻ Guardian
9. How many children do you have?
10. How old are your children?
11. How many times have you visited the emergency department with your children?
  ◻ 1-5 times ◻ 6+times
12. Have any of your children ever been admitted to the hospital?
  ◻ Yes ◻ No

SECTION 2: Assessment of attributes of the arts-based, digital tools, All items were assessed using a 5-point Likert Scale, except items 10 and 11 which allowed open text responses.

1. It is useful.
2. It provides information that is relevant to me as a parent.
3. It is simple to use.
4. I can use it without written instructions or additional help.
5. Its length is appropriate.
6. It is aesthetically pleasing (i.e., images, colours, etc.).
7. It helps me to make decisions about my child’s health.
8. I would use it in the future.
9. I would recommend it to a friend.
10. List the most negative aspects: [open text]
11. List the most positive aspects: [open text]

## Appendix J: KT Tool Development Timeline

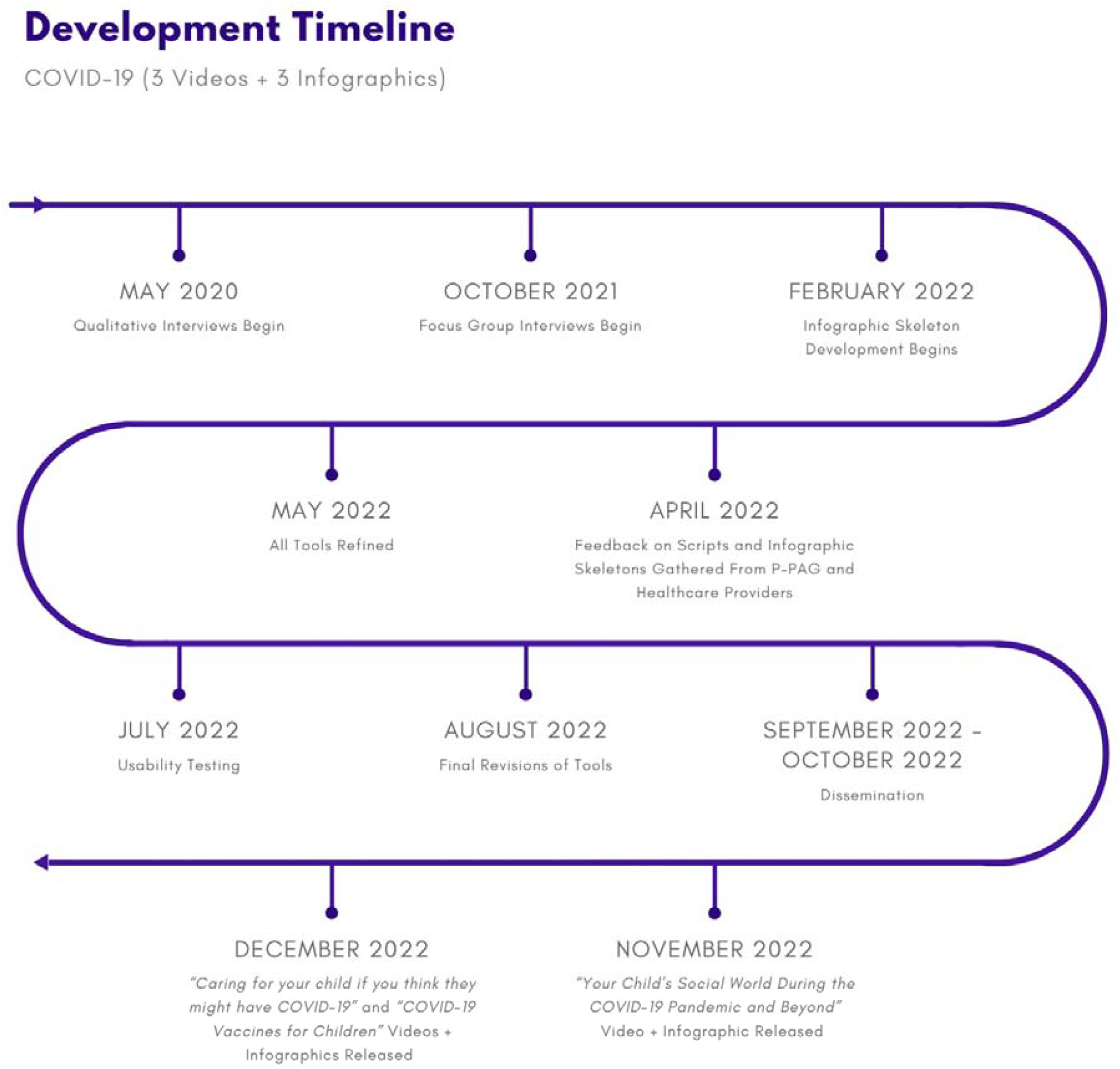

## Research publications

1. Louie-Poon, S., Reid, K., Appiah, P.O., Hartling, L., Scott, S.D. (Published 02 April 2024). “There is a strangeness in this disease”: A qualitative study of parents’ experiences caring for a child diagnosed with COVID-19. PLOS ONE. 10.1371/journal.pone.0300146

2. Cyrkot, S., Hartling, L., Scott, S.D., Elliott, S. (Published 20 March 2024). Parents’ user experience accessing and utilizing a web-based map of Covid-19 recommendations for health decision making: a qualitative study. JMIR Formative Research, 8. 10.2196/53593

3. Bialy, L., Elliott, S.A., Melton, A., Ali, S., Scott, S.D., Knisley, L., Hartling, L. (Published 19 March 2024). Consequences of the Coronavirus disease 2019 pandemic on child and adolescent mental, psychosocial, and physical health: A scoping review and interactive evidence map. Journal of Child Health Care. 10.1177/13674935241238794

4. Elliott, S., Hartling, L., Knisley, L., Wright, K., Scott, S.D. (Published 22 February 2024). “It’s quite a balancing act”: a qualitative study of parents’ experiences and information needs related to the COVID-19 pandemic. Health Expectations, 27(1), e13994. 10.1111/hex.13994

## Conference presentations

1. Cyrkot, S., Hartling, L., Scott, S.D., Elliott, S. Parents’ user experience accessing and utilizing a web-based map of COVID-19 recommendations for health decision making: a qualitative study. Poster presentation. 2024 Pediatric Research Day, University of Alberta. Edmonton, AB. April 17, 2024.

2. Scott, S.D., Hartling, L., Elliott, S.A., Reid, K., Knisley, L. Highlights and challenges of co-creating evidence-based knowledge translation tools for parents about the covid-19 pandemic. Oral presentation. Cochrane Colloquium. London, UK. September 4-6, 2023.

3. Elliott, S.A., Scott S.D., Schunemann, H.J., Hartling, L. Knowledge mobilization of covid-19 evidence-based health recommendations for parents: a multi-methods randomised trial. Poster presentation. Cochrane Colloquium. London, UK. September 4-6, 2023.

4. Scott, S.D., & Hartling, L. Making COVID19 research relatable: Our experience co-developing and evaluating six knowledge translation tools for families. Sigma’s 34th International Nursing Research Congress. Abu Dhabi, United Arab Emirates. July 20-24, 2023.

5. Scott, S.D., & Hartling, L. Co-developing innovative e-tools with parents that merge research and parental experience: a multidisciplinary program of research. Poster presentation. International Council of Nurses (ICN) 2023 Congress. Montréal, QC. July 1-5, 2023.

6. Scott, S.D., & Hartling, L. Co-developing evidence-based knowledge translation tools for parents: Overview of a research program. Child Health and Wellness Seminar [virtual]. Alberta Children’s Hospital Research Institute (ACHRI). January 16, 2023.

7. Wright, K., Elliott, S., Hartling, L., Scott, S.D., Knisley, L. Parenting during the pandemic: a qualitative study of parents’ experience and information needs. Poster presentation. 2022 Northwest SPOR Collaborative Forum. Edmonton/Calgary, AB. October 3-4, 2022.

8. Scott, S.D., Hartling, L. Overcoming emerging COVID19 challenges with relatable science: Developing and evaluating knowledge translation tools for families. Oral presentation (virtual). Sigma’s 33rd International Nursing Research Congress. Edinburgh, Scotland. July 21-25, 2022.

9. Scott, S.D., Hartling, L. Overcoming emerging COVID19 challenges with relatable science: Developing and evaluating knowledge translation tools for families. Poster presentation. KT Canada Scientific Meeting 2022 (virtual). May 4-6, 2022.

10. Bialy, L., Elliott, S.A., Melton, A., Ali, S., Scott, S.D., Hartling, L. Impact of COVID-19 on the unintended physical, psychosocial and mental health effects of children: a living scoping review. Oral presentation. 2022 Pediatric Research Day. Edmonton, AB. April 20, 2022.

11. Ali, S., Elliott, S., Hartling, L., Scott, S.D. Impact of COVID-19 on emergency care for children: a living scoping review. 2022 Pediatric Research Day. Edmonton, AB. April 20, 2022.

## Media coverage

1. Scott, S.D., & Hartling, L. (2022, December 9). Article by: University of Alberta. U of A in the News: More U of A expert, research and institutional news. U of A Leaders’ Morning News Brief.

2. Scott, S.D., & Hartling, L. (2022, December 9). Article by: Henna Saeed. New health tools to answer COVID-19 questions. CityNews Calgary. https://calgary.citynews.ca/2022/12/09/new-health-tools-to-answer-covid-19-questions/amp/

3. Scott, S.D. & Hartling, L. (2022, December 8). Interviewed by Global News at Noon Edmonton. U of A researchers create tools to help parents navigate COVID-19 [Television broadcast]. Global News. https://globalnews.ca/video/9335797/u-of-a-researchers-create-tools-to-help-parents-navigate-covid-19/

4. Scott, S.D. & Hartling, L. (2022, December 8). Interviewed by Henna Saeed. New health tools to answer Albertan’s COVID-19 questions [Television broadcast]. CityNews Calgary. https://calgary.citynews.ca/video/2022/12/08/new-health-tools-to-answer-albertans-covid-19-questions/

5. Scott, S.D. & Hartling, L. (2022, December 8). Article by: University of Alberta. U of A in the News: U of A experts tap parents to create information for parenting with COVID-19. U of A Leaders’ Morning News Brief.

6. Scott, S.D., & Hartling, L. (2022, December 7). Article by: Adam Toy. Alberta experts tap parents to create information for parenting with COVID. Global News. https://globalnews.ca/news/9333676/alberta-coronavirus-update-december-7-2022-parenting/

7. Scott, S.D., & Hartling, L. (2022, December 6). Article by: Chelsea Novak. New research-backed tools ready to support parents in navigating COVID-19 pandemic. Folio, University of Alberta. https://www.ualberta.ca/folio/2022/12/new-research-backed-tools-ready-to-support-parents-in-navigating-covid-19-pandemic.html

8. Scott, S.D. (2021, February 10). Interviewed by Su-Ling Goh. Health Matters: Guidance for parents of kids with COVID-19 [Television broadcast]. Global News. https://globalnews.ca/video/7633828/health-matters-guidance-for-parents-of-kids-with-covid-19

## References

1. Rothan HA, Byrareddy SN. The epidemiology and pathogenesis of coronavirus disease (COVID-19) outbreak. Journal of Autoimmunity. 2020;109:102433.

2. Castagnoli R, Votto M, Licari A, Brambilla I, Bruno R, Perlini S, et al. Severe acute respiratory syndrome coronavirus 2 (SARS-CoV-2) infection in children and adolescents: a systematic review. JAMA pediatrics. 2020;174(9):882–9.

3. Wald ER, Schmit KM, Gusland DY. A pediatric infectious disease perspective on COVID-19. Clinical Infectious Diseases. 2021;72(9):1660–6.

4. Shane AL, Sato AI, Kao C, Adler-Shohet FC, Vora SB, Auletta JJ, et al. A pediatric infectious diseases perspective of severe acute respiratory syndrome coronavirus 2 (SARS-CoV-2) and novel coronavirus disease 2019 (COVID-19) in children. Journal of the Pediatric Infectious Diseases Society. 2020;9(5):596–608.

5. Wu Z, McGoogan JM. Characteristics of and important lessons from the coronavirus disease 2019 (COVID-19) outbreak in China: summary of a report of 72 314 cases from the Chinese Center for Disease Control and Prevention. jama. 2020;323(13):1239–42.

6. Dong Y, Mo X, Hu Y, Qi X, Jiang F, Jiang Z, et al. Epidemiology of COVID-19 among children in China. Pediatrics. 2020;145(6).

7. Shen K-L, Yang Y-H. Diagnosis and treatment of 2019 novel coronavirus infection in children: a pressing issue. World Journal of Pediatrics. 2020;16(3):219–21.

8. Team CC-R, Team CC-R, Team CC-R, Bialek S, Gierke R, Hughes M, et al. Coronavirus disease 2019 in children—United States, february 12–april 2, 2020. Morbidity and Mortality Weekly Report. 2020;69(14):422–6.

9. Rasmussen SA, Thompson LA. Coronavirus disease 2019 and children: what pediatric health care clinicians need to know. JAMA pediatrics. 2020;174(8):743–4.

10. Tagarro A, Epalza C, Santos M, Sanz-Santaeufemia FJ, Otheo E, Moraleda C, et al. Screening and severity of coronavirus disease 2019 (COVID-19) in children in Madrid, Spain. JAMA pediatrics. 2021;175(3):316–7.

11. Scarpellini F, Segre G, Cartabia M, Zanetti M, Campi R, Clavenna A, et al. Distance learning in Italian primary and middle school children during the COVID-19 pandemic: a national survey. BMC public health. 2021;21(1):1035.

12. Cost KT, Crosbie J, Anagnostou E, Birken CS, Charach A, Monga S, et al. Mostly worse, occasionally better: impact of COVID-19 pandemic on the mental health of Canadian children and adolescents. European child & adolescent psychiatry. 2021:1-

13. Golberstein E, Wen H, Miller BF. Coronavirus disease 2019 (COVID-19) and mental health for children and adolescents. JAMA pediatrics. 2020;174(9):819–20.

14. McKinnon B, Quach C, Dubé È, Nguyen CT, Zinszer K. Social inequalities in COVID-19 vaccine acceptance and uptake for children and adolescents in Montreal, Canada. Vaccine. 2021;39(49):7140–5.

15. Larivière-Bastien D, Aubuchon O, Blondin A, Dupont D, Libenstein J, Séguin F, et al. Children’s perspectives on friendships and socialization during the COVID-19 pandemic: A qualitative approach. Child: Care, Health and Development. 2022;48(6):1017–30.

16. Vaillancourt T, Beauchamp M, Brown C, Buffone P, Comeau J, Davies S, et al., editors. Children and schools during COVID-19 and beyond: Engagement and connection through opportunity. Royal Society of Canada; 2021.

17. Alteri C, Scutari R, Costabile V, Colagrossi L, Yu La Rosa K, Agolini E, et al. Epidemiological characterization of SARS-CoV-2 variants in children over the four COVID-19 waves and correlation with clinical presentation. Scientific reports. 2022;12(1):10194.

18. Lee BR, Harrison CJ, Myers AL, Jackson MA, Selvarangan R. Differences in pediatric SARS-CoV-2 symptomology and Co-infection rates among COVID-19 Pandemic waves. Journal of Clinical Virology. 2022;154:105220.

19. Hartling L, Elliott SA, Buckreus K, Leung J, Scott SD. Development and evaluation of a parent advisory group to inform a research program for knowledge translation in child health. Research Involvement and Engagement. 2021;7(1):38.

20. Hartling L, Elliott SA, Mabbott A, Leung J, Shearer K, Smith C, et al. Four year evaluation of a parent advisory group to support a research program for knowledge translation in child health. Research Involvement and Engagement. 2024;10(1):14.

21. Louie-Poon S, Reid K, Appiah PO, Hartling L, Scott SD. “There is a strangeness in this disease”: A qualitative study of parents’ experiences caring for a child diagnosed with COVID-19. PloS one. 2024;19(4):e0300146.

22. Hartling L, Elliott SA, Wright KS, Knisley L, Scott SD. ‘It’s quite a balancing act’: A qualitative study of parents’ experiences and information needs related to the COVID-19 pandemic. Health Expectations. 2024;27(1):e13994.

